# Environmental Chemicals as Modifiers of the Association between Chronological Age and Ovarian Reserve

**DOI:** 10.64898/2025.12.09.25341902

**Authors:** Ashley I Naimi, Edward H. Kennedy, Ya-Hui Yu, Russ Hauser, Lidia Minguez-Alarcon, Audrey Gaskins

## Abstract

**Objective:** To evaluate whether the well-established age-related reduction in antral follicle counts (AFC) is greater among women with higher concentrations of endocrine-disrupting chemicals.

**Design:** Prospective cohort study using the doubly robust (DR) learner, a flexible machine learning method that models the relationship between an exposure and outcome of interest and identifies potential modifiers of this relation.

**Subjects:** Seven hundred seventy-five women aged 18-45 years enrolled in The Environment and Reproductive Health (EARTH) Study at the Massachusetts General Hospital Fertility Center, Boston, USA, between 2004 and 2019.

**Exposure:** Age at AFC assessment calculated as the date of antral follicle scan minus the patient’s birthdate, categorized into <35 years or ≥35 years. We assessed urinary concentrations of 16 endocrine-disrupting chemicals, including 11 urinary phthalate metabolites, 3 urinary parabens, urinary bisphenol A, and hair mercury, as potential modifiers of the age-AFC association.

**Main Outcome Measures:** AFC assessed by transvaginal ultrasonography.

**Results:** In this cohort of 775 women, 43% were less than 35 years of age. Women greater than or equal to aged 35 years had lower AFC (mean [SD]: 11.63 [6.24] vs. 16.89 [8.64]) Four endocrine-disrupting chemicals significantly modified the age-AFC association. Women with higher urinary mono-isobutyl phthalate (MiBP), monocarboxyisooctyl phthalate (MCOP), and mono(2-ethylhexyl) phthalate (MEHP) concentrations experienced stronger negative associations between age and AFC. Analyses revealed that the age-AFC association strengthened, declining at a faster rate when MiBP, MCOP, or MEHP concentrations were higher. Women with higher hair Hg concentrations experienced weaker negative associations between age and AFC. The remaining 12 endocrine-disrupting chemicals did not modify the age-AFC relationship.

**Conclusion:** Select phthalate metabolites, specifically MiBP, MCOP, and MEHP, were associated with a modified age-related decline in ovarian reserve in a dose-dependent manner. These findings, though limited, add to the growing evidence that certain phthalates are associated with a steeper age-related decline in AFC.

**What this study adds:** Using the DR-Learner, a flexible machine learning approach designed to identify effect modifiers among a large set of candidates, we evaluated whether 16 endocrine-disrupting chemicals modify the well-established association between age and ovarian reserve. Three phthalate metabolites (MiBP, MCOP, MEHP) strengthened the negative age-antral follicle count association; for MiBP and MCOP this pattern was curvilinear and dose-dependent, with apparent concentration thresholds above which the age-related decline accelerated. Hair mercury was associated with a weaker negative association, possibly reflecting protective effects of seafood-derived omega-3 fatty acids. We discuss important limitations, but note that our findings add to evidence that higher phthalate concentrations accompany a stronger negative age–AFC association.

## Introduction

Chronological age is the strongest predictor of ovarian reserve,^1^ with reductions in the number of antral follicles (a measure of reproductive capacity) negatively related to increasing age.^2^ Several recent studies have demonstrated that a range of environmental endocrine distrupting chemicals (EDCs) may alter the rate at which ovarian reserve is depleted with age.^3–6^ EDCs are thought to hasten this loss through several biological pathways, including oxidative stress, disruption of ovarian steroidogenesis, and premature activation and atresia of ovarian follicles.^6^ Understanding how environmental chemicals may modify the association between age and ovarian reserve could offer important insights into the mechanisms of reproductive aging. If certain exposures are found to accelerate depletion, interventions to reduce these exposures may help preserve ovarian reserve and improve reproductive outcomes.

However, many environmental EDCs exist. This leads to a potentially large degree of complexity between the constellation of chemicals, and the effects these may have on the relation between chronological age and ovarian reserve.^7^ For example, higher order interactions between a subset of EDCs may result in greater than additive reductions in the rate at which ovarian reserve is depleted with age.^8^ Thus, while this provides public health opportunities to efficiently and effectively reduce the negative impact of EDCs on fertility, commonly used regression based methods are ill-suited to the task of uncovering these scenarios.^9^

Recently, several advances have been made in using machine learning to model and evaluate effect measure modification that better accomodate multiple effect modifiers and their interactions. These methods include the doubly robust (DR) Learner,^10^ developed to flexibly model the relationship between an exposure and outcome of interest, and identify potential modifiers of this relation among a set of candidate modifiers. Here, we use the DR-Learner to evaluate whether the well-known age-related reduction in antral follicle count (AFC) is greater among women with higher concentrations of EDCs in 775 women seeking fertility care at an academic medical center.^11^

## Methods

### Study Population

The Environment and Reproductive Health (EARTH) Study was an observational cohort study (2004-2019) that recruited women aged 18 to 45 years (and their partners) who were presenting for fertility treatment and evaluation at the Massachusetts General Hospital (MGH) Fertility Center in Boston, MA.^11^ Approximately 60% of eligible women contacted by the research staff participated in the study. Women could enroll in the EARTH Study at any point during their care at MGH, including at the start of their fertility investigation or after initiating treatment. Data were collected primarily to evaluate how exposure to environmental chemicals and lifestyle factors affect reproductive health. This analysis included women who provided at least one hair or urine sample for the measurement of environmental chemicals and had at least one measurement of AFC at the MGH Fertility Center.

Of the initial 1022 antral follicle scans (from 869 women) that were available for analysis, we excluded 42 that were done while the woman was on Lupron, 21 incomplete scans, 77 scans done on women with polycystic ovaries, and 107 repeated scans. Thus, our final sample size included 775 women in EARTH that contributed one measurement of ovarian reserve and had at least one environmental chemical measured. The study was approved by the Human Studies Institutional Review Boards of the MGH, the Harvard T.H. Chan School of Public Health, and the Centers for Disease Control and Prevention (CDC). Participants signed an informed consent after the study procedures were explained by trained research study staff and all questions were answered.

### Exposure

Age at AFC assessment was calculated as the date of antral follicle scan minus the patient’s birthdate, rounded down to the nearest integer. We categorized patient age into <35 years or ≥35 years, to align with the American College of Obstetricians and Gynecologists (ACOG) definition of advanced maternal age.^12^

### Outcome

Ovarian AFC, defined as an integer variable of the sum of antral follicles in both ovaries, was measured by a reproductive endocrinologist using transvaginal ultrasonography on the 3rd day of an unstimulated menstrual cycle or on the 3rd day of a progesterone withdrawal bleed. No fertility medications were used in the cycle prior to the antral follicle scan. AFC scan values were not truncated or trimmed.

### Covariates: Environmental Chemicals

The EARTH Study measured 22 urinary EDCs, including phthalate metabolites, phenols, parabens, organophosphate flame retardant (OPFR) metabolites, as well as hair mercury concentrations. Missing data exceeded 40% in seven chemicals (bisphenol S, bisphenol F, benzophenone-3, triclosan, and three OPFR metabolites), which precluded their inclusion in the current analysis. We assessed urinary concentrations of eleven phthalate metabolites [mono-n-butyl phthalate (MBP), mono-isobutyl phthalate (MiBP), monocarboxyisononyl phthalate (MCNP), monocarboxyisooctyl phthalate (MCOP), mono(2-ethyl-5-carboxypentyl) phthalate (MECPP), mono(2-ethyl-5-hydroxyhexyl) phthalate (MEHHP), mono(2-ethylhexyl) phthalate (MEHP), mono(2-ethyl-5-oxohexyl) phthalate (MEOHP), mono(3-carboxypropyl) phthalate (MCPP), monoethyl phthalate (MEP), monobenzyl phthalate (MBzP), three parabens [methylparaben (MP), propylparaben (PP), and butylparaben (BP)], and one phenol [bisphenol A (BPA)]. We also evaluated hair mercury (Hg) concentrations, a heavy metal.

In brief, EARTH participants provided a urine sample at study entry and up to twice during a subsequent treatment cycle using a sterile polypropylene cup. We selected the urine sample that was provided closest in time to AFC assessment for this analysis. Upon collection, each urine sample was analyzed for specific gravity (SG) with a handheld refractometer (National Instrument Company, Inc., Baltimore, MD, USA), divided into aliquots, and frozen at −80°C. Urine samples were later shipped on dry ice overnight to the CDC (Atlanta, GA, USA) for quantification of the urinary chemicals. Urinary phthalate, paraben, and phenol concentrations were measured using online solid-phase extraction–high performance liquid chromatography–isotope dilution tandem mass spectrometry, as previously reported.^13–15^ Standard QA/QC procedures were followed for each analysis. Because samples were analyzed across multiple batches over the study period (2004–2019), the LOD for a given analyte varied across batches owing to differences in reagent lots, instrument calibration, and periodic method refinements; Table S1 reports the range of LODs for each chemical. Urinary concentrations below the LOD were replaced with the LOD divided by the square root of two.^16^ We corrected the urinary concentrations for SG using the following formula: Pc = P[(1.016 – 1)/SG – 1], where Pc is the SG-corrected concentration, P is the measured concentration, and 1.016 is the median SG level in the study population. We used SG-corrected urinary concentrations in all analyses.

We also assessed hair mercury (Hg) levels in EARTH women. Hair samples were collected in person, or at home and mailed to study staff in an envelope at study entry. Study participants were instructed to collect hair samples on clean hair. The hair sample was also cleaned before analysis by sonication for 15 minutes in a 1% Triton X-100 solution before analysis to remove extraneous contaminants. Samples were then rinsed with distilled deionized water and dried 5 times at 60°C for 48 hours. Total Hg in nanograms per gram (ng/g) was measured using 0.02 g of the proximal 2 cm of hair using a Direct Mercury Analyzer 80 (Milestone Inc, Monroe, CT) with a matrix matched calibration curve. We used certified reference material GBW 07601 (human hair; Institute of Geophysical and Geochemical Exploration, China) containing 360 ng/g mercury as the quality control standard.

### Covariates: participants’ characteristics

Weight and height were measured at study entry by trained study staff. Body mass index (BMI) was calculated as weight (in kilograms) per height (in meters) squared. The detailed take-home questionnaire contained questions on demographics (e.g. race, education), lifestyle factors (e.g. smoking history), reproductive health (e.g. menstrual cycle characteristics, infertility treatment history), and medical history (e.g. diagnosis of major chronic diseases). Clinical information, such as infertility diagnosis and parity/gravidity were abstracted from the patient’s electronic medical records by trained study staff.

As noted, variables with more than 40% missing data were excluded from the analyses. The remaining variables had no more than 18% missingness, and were imputed via a random forest algorithm for missing data,^17^ with 50 iterations and 2,000 trees. In the models used to construct our DR-learner, we also included variables that served as indicators of whether a particular value was imputed or not, to add flexibility to our nuisance function estimation. Details of missing data analyses and imputation are available in the GitHub Repository. The final set of covariates (*X*) used in the analyses included year and month of AFC measurement, smoking status (never, current, former), race, education, BMI, previous intrauterine insemination, previous in vitro fertilization, gravidity, and sixteen environmental chemicals noted above.

### Statistical Analysis

#### Estimand

We are interested in two key covariate-adjusted associations between age (denoted A) and AFC (denoted Y). First, we define

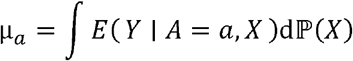

as the standardized average AFC among those aged <35 years (*a* = 0) or ≥ 35 years (*a* =1). Thus, µ_*a*_ represents the age-specific mean AFC standardized to the overall distribution of all covariates *X*. Using this, we define our first parameter of interest as

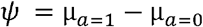

which represents the mean difference in AFC standardized to the distribution of all covariates *x* in EARTH. This can be viewed as the difference in mean counts for older versus younger participants, whose covariate distributions were otherwise the same.

Our second parameter of interest represents the *X* = *x* specific association between age and antral follicle count, denoted as

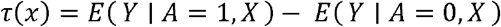

This represents the mean difference in AFC among older versus younger participants with covariate values *X* = *x*. For example, if we let *x* denote specific concentrations of hair Hg (in ng/g) and BPA (in μg/L), we can estimate the association between age and AFC for women with these concentrations. If *τ*(*x*) varies across values of *x*, then modification of the association by *x* is present.

#### Doubly Robust Learner (DR-Learner)

The *DR-Learner* begins with predictions from the a supervised machine learning fit for the propensity score [here denoted 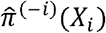] and outcome model [here denoted 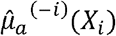] for each individual in the sample. These predictions are then combined to obtain estimates of a doubly robust pseudo-outcome for each individual in the sample, according to the following formula:

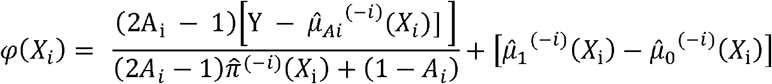

where 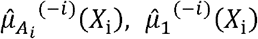 and 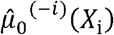are predictions from the outcome model under *A*_*i*_, *A*_*i*_ = 1 and *A*_*i*_ = 0, respectively. Here, and throughout, the superscript − *i* notation denotes estimates obtained in out-of-sample cross-validation folds.^18,19^ For example, 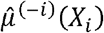 denotes predictions for individual *i* from a model 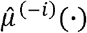that was fit to a subset of the data that does not include individual *i* . This pseudo-outcome, which we refer to as the augmented inverse probability weighted (AIPW) score, is the efficient influence function for the standardized mean difference parameter *Ψ* . We can estimate *Ψ* by taking the average of this AIPW score.^10^ However, to estimate the *x* specific standardized mean difference *Ψ* (*x*), we regress the individual AIPW scores against the set of covariates *X* to capture how the mean difference between age and AFC changes as a function of these variables. Importantly, unlike other CATE learners,^20^ the DR-learner enables us to regress the AIPW scores against a subset of the variables in *X* (i.e., only the environmental chemicals, excluding the participant characteristics). However, to avoid introducing additional notation (and thus slightly abusing the established notation), we refer to these environmental chemical variables throughout collectively as *X*.

We used the 10-fold cross-validated (CV) Super Learner to estimate 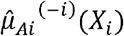 and 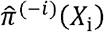. The same set of covariates *X* (environmental chemicals and participant characteristics) was used to model both propensity score and outcomes. The Super Learner library included random forest,^21^ penalized regression,^22^ extreme gradient boosting,^23^ multivariate adaptive regression splines,^24^ the simple mean estimator and generalized linear models.^25^ Where applicable, variations of each of these algorithms were also included in the stacking algorithm, to allow for differing tuning parameters for eligible algorithms.^26^ We also included in the stacking algorithm a correlation-based screening procedure to a generalized linear model with all two way interactions, which selected only those interactions that contribute to the fit. Additional details on the specific set of tuning parameters and procedure of deploying screening procedures included in the stacking algorithm are available in the associated online web supplement and GitHub repository.

#### Summary of conditional associations between age and AFC

To identify which environmental chemicals are important modifiers of the association between age and AFC, we used a “best linear projection” modeling approach which can be implemented using linear regression to regress the AIPW score on environmental chemicals. We construct a linear projection fit, denoted *α X*, where *α* denotes (*p* +1) ×1 vector of regression coefficients, and *X* is an *n* × (*p* + 1) design matrix of *p* log-transformed EDC measurements along with a column of 1s for the intercept. We then estimate the coefficients by solving:

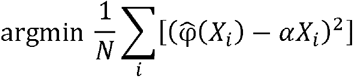

In this case *α*_*j*_ captures the extent of which the environmental chemical *X*_*j*_ linearly explains variation in the mean difference of AFC comparing ages ≥35 years vs. < 35 years across observations in the sample, conditional on all other EDCs in the model. Alternatively, we can obtain *unconditional* measures of the extent to which the EDCs in *X* modify the assocaiton between age and AFC by fitting a linear model with only a single variable, in which case *αX* includes on the intercept and a term for the EDC of interest.

This linear projection approach provides a simple way to summarize the relationship between the standardized mean difference and the covariates as a linear function, instead of capturing the exact relationships.^27^ Therefore, after our linear projection modeling analyses, we explored how the difference in mean AFC between women aged ≥35 years and those <35 years changes with each EDC by using a Super Learner algorithm that included a combination of generalized additive models and the locally estimated scatterplot smoothing (LOESS) algorithm, which allowed us to explore the potential curvilinear relationships between the standardized mean difference as a function of specific the EDCs.^28^

## Results

In this cohort of 775 women, 43% were less than 35 years of age. Compared to those aged less than 35 years, women greater than or equal to aged 35 years were more likely to have higher BMI, gravidity greater than zero, and history of IUI or IVF. They also had lower AFC (mean [SD]: 11.63 [6.24] vs. 16.89 [8.64]). These two groups also had different distributions of key environmental chemicals (Table 1, Table S1). Table S2 presents the unconditional and conditional assocaitions between each EDC and AFC from models stratified by age. Table 1 presents summary statistics for the distributions of all 16 environmental variables, and shows that most are highly skewed to the right, suggesting the benefit of log-transforming them in the linear projection models. The median (Q1, Q3) time from the AFC scan date to the measurement of EDCs excluding Hg was four weeks (0.6, 12). The median (Q1, Q3) time from the AFC scan date to the measurement of Hg was 4.4 weeks (0, 14).

**Table 1.**
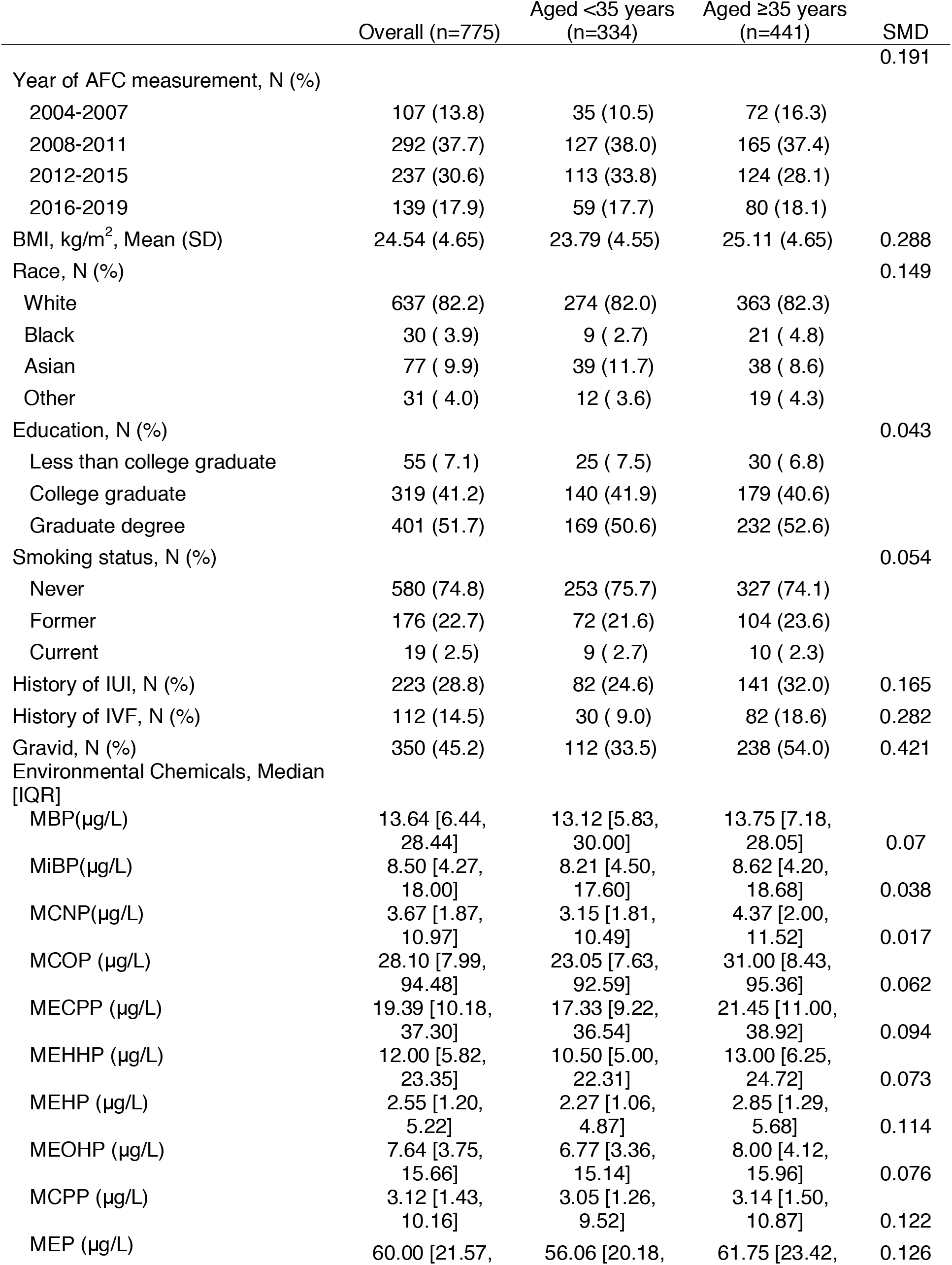
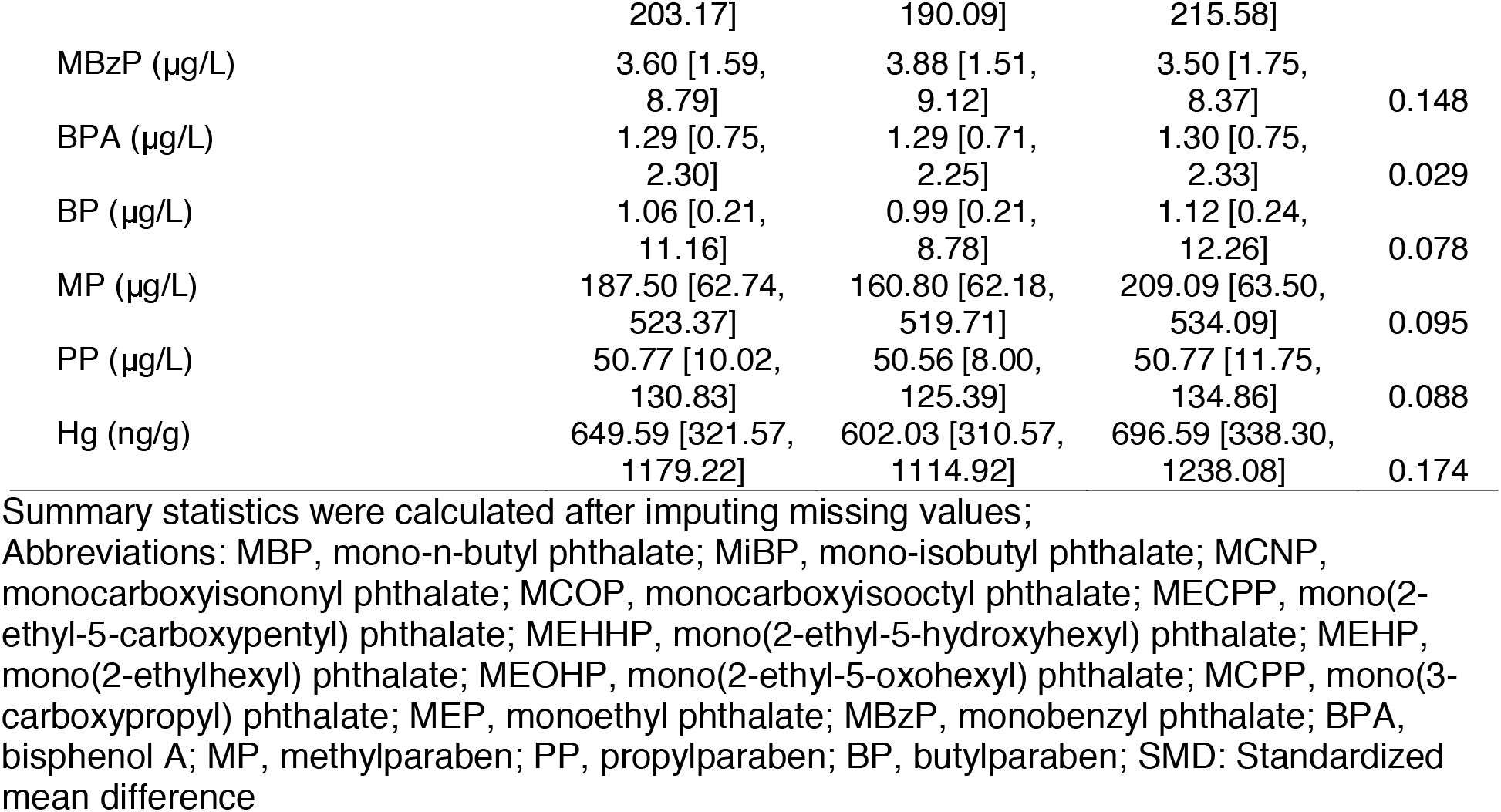
Characteristics of 775 women, overall and by age groups in the EARTH Study, 2004-2019.

### Overall and Best Linear Projection Model Results

We estimated a standardized mean difference in AFC of -5.1 (95% confidence intervals: -6.2, -3.9), which suggests that, on average and adjusting for all other observed covariates women ≥35 years had five fewer antral follicles than women <35 years. Figure 1 shows a plot of the Wald test statistics (coefficient / standard error) from the conditional and unconditional linear projection models which regressed the AIPW scores obtained from the DR-Learner on all 16 log-transformed environmental chemicals. The four environmental chemicals with the largest test statistics were hair Hg, as well as urinary MiBP, MCOP, and MEHP. The direction of the test statistic for hair Hg was positive, while the direction for MiBP, MCOP, and MEHP were negative, suggesting that higher concentrations of hair Hg attenuate (improve) the negative association between age and AFC, while higher concentrations of MiBP, MCOP, and MEHP increase (worsen) the negative association between age and AFC.

**Figure 1.**
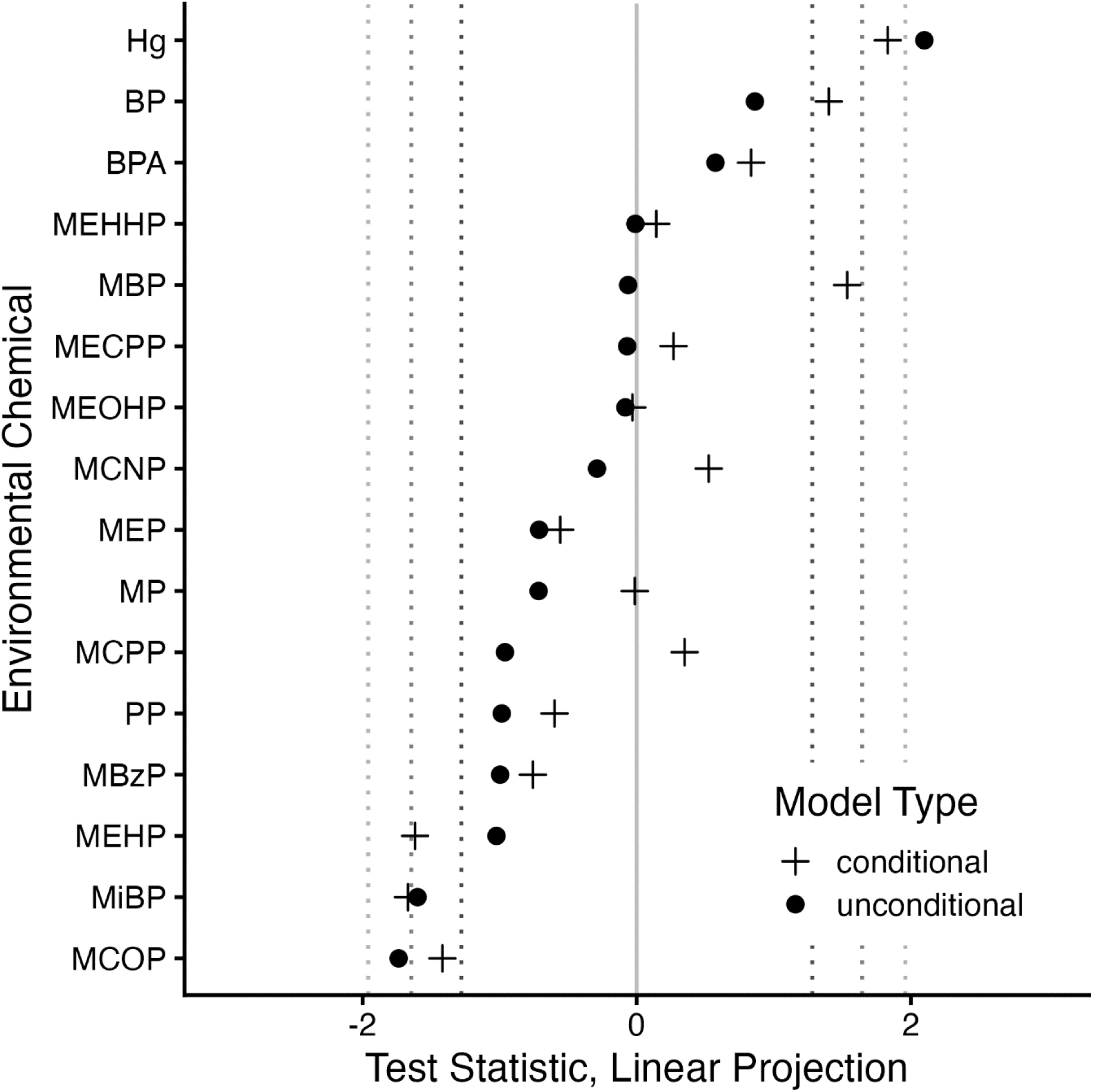
Scatterplot of Wald test statistics from the linear projection models that regress the AIPW scores constructed with the DR-Learner against the log-transformed EDC variables included in our analysis from 775 women in the EARTH Study, 2004-2019. This Figure plots points from conditional (coefficients estimated with all EDC variables in the model, represented as filled circles) and unconditional (coefficients estimated with only one EDC variable in the model, represented as circles) linear projection models. The dashed vertical lines represents critical Wald test statistic values of |1.28| (darkest), |1.44|, and |1.64| (lightest), representing two-sided α-levels of roughly 20%, 15%, and 10%, respectively.

Because they were identified as playing the largest role in the linear projection models, we further explored the role of MiBP, MCOP, and MEHP as effect modifiers by using a cross-validated locally estimated scatterplot smoother (cvLOESS) to summarize the mean difference across the range of these three environmental chemicals (additional results for all environmental chemicals is provided in the online GitHub Repository). Figure 2 demonstrates how these relationships change in our data, with two of the three suggesting important threshold effects.

**Figure 2.**
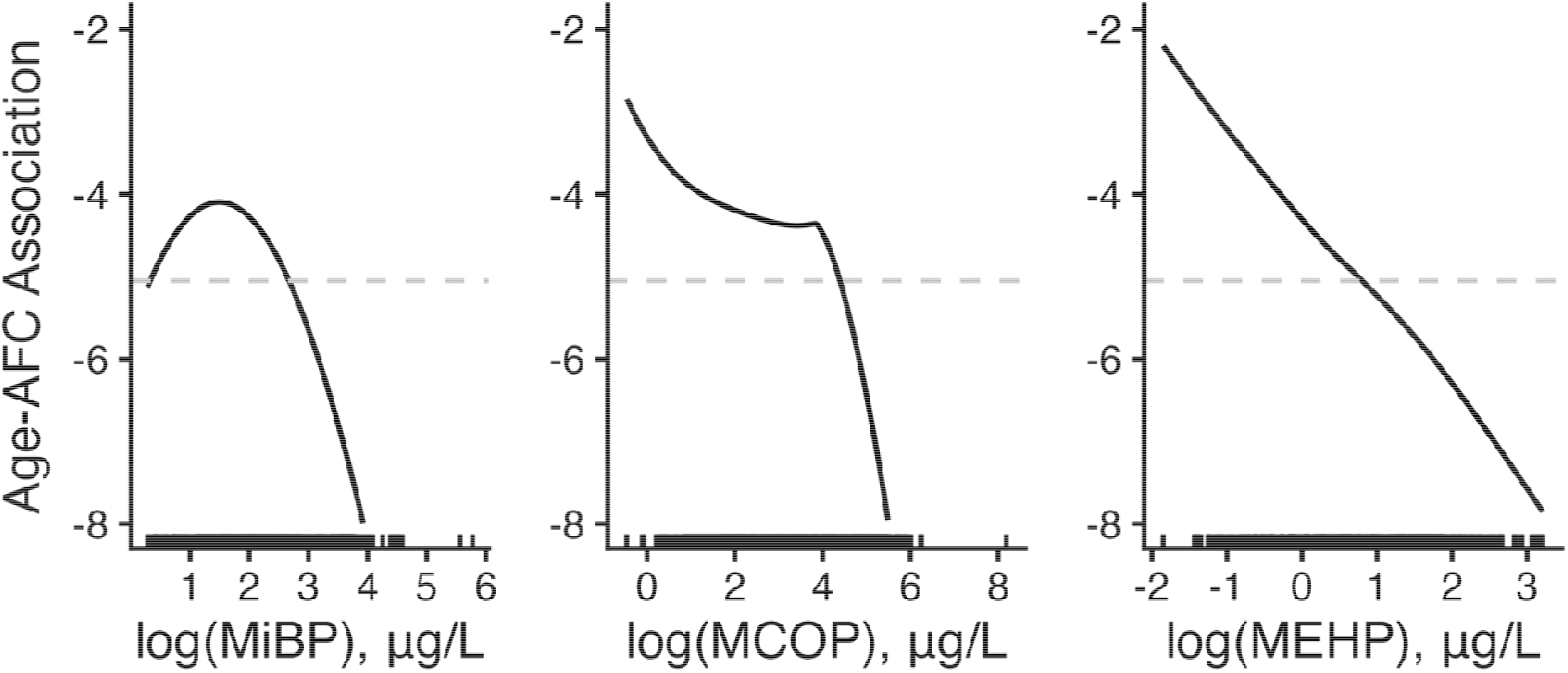
Conditional mean difference of AFC comparing women aged ≥35 years vs. < 35 years across MiBP, MCOP, and MEHP concentrations obtained from the DR-Learner with 775 women in the EARTH Study, 2004-2019. Rug-plots demonstrating the distribution of women in the sample across the domain of each modifier. Concentrations are adjusted for urinary specific gravity and log-transformed. The gray dashed line represents the overall association between age and AFC.

For example, for women with specific-gravity–adjusted MiBP concentrations beyond ≈ 4.5 μg/L (natural-log scale ≈ 1.5, roughly the 25th percentile of the MiBP distribution), the negative difference in mean AFC between women ≥35 versus <35 years increased in magnitude. A similar pattern was observed for MCOP concentrations beyond ≈ 50 μg/L (log scale ≈ 4, roughly the 60th percentile of the MCOP distribution). In contrast, the negative difference in mean AFC between women ≥35 versus <35 years increased steadily across the full range of MEHP concentrations, with no apparent threshold (Figure 2).

## Discussion

In this cohort of 775 women seeking fertility evaluation and treatment, we identified four environmental chemicals or their metabolites that played a moderately strong role in modifying the association between age and AFC. Our use of flexible machine learning methods allowed us to identify potentially important modifiers among 16 EDCs while accounting for their complex relationships, an area where tranditional parametric regression is often limited. In addition, we estimated dose-response relationships for the association between age and antral follicle count with two phthalate metabolites [MiBP and MCOP], which showed curvilinear patterns and apparent thresholds above which the negative age–AFC association was more pronounced. These findings add to the growing literature on the potentially deleterious effects of phthalates on female reproduction.

Women with higher hair mercury concentrations had a smaller negative association between age and AFC, which may suggest that high hair mercury attenuates the age-related decline in ovarian reserve. However, this finding should be interpreted carefully. Hair mercury concentrations, particularly in our population of women undergoing fertility treatment, are likely also a biomarker of seafood and long-chain omega-3 fatty acid intake,^4,29^ the latter of which has been shown to yield beneficial effects on reproductive outcomes in the EARTH Study^30^ as well as others.^31–33^ Consequently, the reduced negative association between age and AFC observed with higher hair mercury concentrations may be a proxy for increased consumption of long-chain omega-3 fatty acids from seafood rather than a direct effect of mercury exposure.

In contrast, we found that women with higher concentrations of MiBP, MCOP, and MEHP had stronger negative associations between age and AFC. These chemicals are phthalate metabolites derived from plasticizers and personal care products,^34^ and are increasingly being implicated in altered reproductive function in females.^34–36^ Several biological mechanisms may account for the modifying effects we observed. Experimental and animal models suggest phthalates including di(2-ethylhexyl) phthalate (the parent diester of MEHP) induce oxidative stress in granulosa cells and oocytes, accelerate the activation and subsequent atresia of ovarian follicles, and suppress ovarian steroidogenesis, in part through inhibition of aromatase and altered expression of steroidogenic enzymes.^34,37^ These processes could deplete the follicle pool more rapidly, steepening the decline in AFC across age as we observed. However, this evidence is drawn largely from in vitro and rodent studies, and thus, while consistent with our findings, largely speculative. Further analysis revealed important dose-threshold relationships. Specifically, the association between age and AFC strengthened as MiBP increased beyond ≈ 4.5 μg/L and as MCOP increased beyond ≈ 50 μg/L, suggesting potential thresholds above which these phthalates may accelerate age-related ovarian reserve decline. We expected to see a potential detrimental association between exposure to Hg and ovarian reserve as it has been linked with higher incidence of infertility, menstrual cycle disturbances, and hormonal disorders in women. Experimental models have also shown that Hg exposures affect oogenesis and folliculogenesis in animals. However, most of the detrimental effects of Hg exposure on reproductive function in women have been observed in occupational settings (which differs from exposure to methyl Hg (primarily through fish consumption). The results from animal models are also difficult to extrapolate to humans because of the physiological differences among species, exposure periods and doses, which all lead to variation in observed effects.^38–40^

Our results can be situated within a small and inconsistent literature on phthalates and ovarian reserve. In an earlier analysis of a subset of the EARTH sample, higher urinary DEHP metabolite concentrations (including MEHP) were associated with lower AFC, whereas non-DEHP metabolites such as MiBP and MCOP showed no consistent association.^41^ Our finding that higher MEHP accompanies a steeper age-related decline in AFC is broadly consistent with that earlier work, while the modifying roles we observed for MiBP and MCOP have not, to our knowledge, been reported previously. Beyond this, the evidence is mixed. A subsequent EARTH mixtures analysis found no association between phthalate biomarkers and AFC,^42^ a recent cohort of women undergoing IVF reported higher AFC at greater phthalate exposure (interpreted as accelerated primordial follicle recruitment),^43^ and studies of anti-Müllerian hormone have been inconsistent, with null findings in fertility-clinic populations.^44,45^ Our analysis extends this literature by asking a distinct question: whether these chemicals modify the age– AFC association.

These findings should be considered in light of several limitations. Notable among these is that our data are cross-sectional; many of the EDC measurements occurred after AFC scans were conducted. This limits our ability to make statements about any causal relationships in these data, including the relationship between age and AFC (given, again, cross-sectional data, as well as the chance that some of our models may have conditioned on intermediates) or the role that EDCs play in altering the relation between age and AFC. Thus, while the causal identification conditions usually invoked for estimating average treatment effects^46^ are difficult to justify in this work for a number of reasons (e.g., age is a non-modifiable exposure), causal inference was not, nor could not, be our aim in this work, as many of our exposure measurements came after the AFC scan. Misclassification is also possible, particularly for urinary metabolites: because these non-persistent chemicals have short half-lives and episodic exposure patterns, their concentrations can vary within individuals over time, so the single sample available for each participant may not represent her usual exposure.^47^ Relatedly, concentrations below the limit of detection were assigned the LOD divided by √2, and hair mercury may be subject to additional misclassification. While AFC is a well-accepted measure of ovarian reserve, it is not a perfect surrogate for female fertility potential, which is the underlying construct of interest. Our analysis focused on subfertile women seeking treatment for infertility. Because these women tend to be older, White, and of higher socioeconomic status, the generalizability of our results to all reproductive aged women may be limited. Previous work suggests that infertile women <40 years have similar AFCs compared to age-matched women with no history of infertility, suggesting that our outcome should not be strongly affected by our choice of study population. EARTH participants may also represent a subset of the general population of potentially greater interest given the fertility implications of chronological age. Urinary phthalate metabolite concentrations in our cohort were broadly comparable in magnitude to those reported for reproductive-aged women in the U.S. general population (National Health and Nutrition Examination Survey [NHANES]).^48^ These comparisons are approximate, however, as our values are specific-gravity corrected medians whereas NHANES concentrations are typically reported as uncorrected geometric means. Finally, although dichotomizing age necessarily simplifies its continuous relationship with AFC, we chose 35 years, the American College of Obstetricians and Gynecologists definition of advanced maternal age.^12^ We selected this clinically established, pre-specified cutpoint as it is the most relevant threshold for reproductive research on fertility.

In terms of strengths, data in EARTH allowed us to leverage high-quality information from a well-characterized cohort with detailed exposure assessment and reproductive endpoints that are often unobservable in populations not seeking fertility treatment. By applying the DR-Learner, we were able to identify nonlinear, threshold-dependent relationships that would have been missed by conventional linear models. Our findings suggest that certain phthalates may accelerate age-related declines in ovarian reserve, with potential implications for fertility and reproductive lifespan. Moreover, the dose-threshold patterns we observed for MiBP and MCOP— chemicals ubiquitous in consumer products —highlight the need for further research to confirm these thresholds and inform potential regulatory action.

In conclusion, we found that women with higher concentrations of select phthalate metabolites had greater negative associations between age and ovarian reserve, with evidence of thresholds that warrant further investigation. These findings underscore the importance of considering environmental chemical exposures as modifiable risk factors for reproductive aging and suggest that reducing exposures to certain phthalates could help preserve ovarian function.

## Data Availability

The study investigators have copies of the entire database but cannot release that version to outside investigators due to the permissions granted by the participants during the consent process and to protect participant confidentiality.

---

**Table S1.** Summary statistics of environmental chemicals in the EARTH Study, 2004-2019.

| Chemical | Unit | LOD | Minimum | Maximum | Median | Q25 | Q75 | Skewness | % Missing |
| --- | --- | --- | --- | --- | --- | --- | --- | --- | --- |
| MBP | µg/L | 0.4–0.6 | 0.42 | 315.00 | 13.64 | 6.44 | 28.44 | 4.79 | 12% |
| MiBP | µg/L | 0.2–0.3 | 0.17 | 321.87 | 8.50 | 4.27 | 18.00 | 7.58 | 12% |
| MCNP | µg/L | 0.2–0.6 | 0.19 | 1633.33 | 3.67 | 1.87 | 10.97 | 15.34 | 18% |
| MCOP | µg/L | 0.2–0.7 | 0.62 | 3585.00 | 28.10 | 7.99 | 94.48 | 16.59 | 18% |
| MECPP | µg/L | 0.2–0.6 | 1.20 | 2109.00 | 19.39 | 10.18 | 37.30 | 8.21 | 12% |
| MEHHP | µg/L | 0.2–0.7 | 0.35 | 1977.00 | 12.00 | 5.82 | 23.35 | 11.91 | 12% |
| MEHP | µg/L | 0.5–1.2 | 0.15 | 267.19 | 2.55 | 1.20 | 5.22 | 8.41 | 12% |
| MEOHP | µg/L | 0.2–0.7 | 0.21 | 1278.00 | 7.64 | 3.75 | 15.66 | 12.54 | 12% |
| MCPP | µg/L | 0.1–0.2 | 0.11 | 757.50 | 3.12 | 1.43 | 10.16 | 15.31 | 12% |
| MEP | µg/L | 0.4–0.8 | 2.38 | 7650.00 | 60.00 | 21.57 | 203.17 | 9.38 | 12% |
| MBzP | µg/L | 0.2–0.3 | 0.15 | 189.60 | 3.60 | 1.59 | 8.79 | 7.77 | 12% |
| BPA | µg/L | 0.4 | 0.06 | 85.87 | 1.29 | 0.75 | 2.30 | 11.76 | 12% |
| BP | µg/L | 0.2 | 0.03 | 160.83 | 1.06 | 0.21 | 11.16 | 4.12 | 15% |
| MP | µg/L | 1.0 | 0.39 | 23196.00 | 187.50 | 62.74 | 523.37 | 18.96 | 15% |
| PP | µg/L | 0.2 | 0.04 | 4185.00 | 50.77 | 10.02 | 130.83 | 12.58 | 15% |
| Hg | ppm | 0.01 | 2.74 | 8598.12 | 649.59 | 321.57 | 1179.22 | 2.60 | 11% |

Summary statistics were calculated after imputing missing values;
Abbreviations: MBP, mono-n-butyl phthalate; MiBP, mono-isobutyl phthalate; MCNP, monocarboxyisononyl phthalate; MCOP, monocarboxyisooctyl phthalate; MECPP, mono(2-ethyl-5-carboxypentyl) phthalate; MEHHP, mono(2-ethyl-5-hydroxyhexyl) phthalate; MEHP, mono(2-ethylhexyl) phthalate; MEOHP, mono(2-ethyl-5-oxohexyl) phthalate; MCP, mono(3-carboxypropyl) phthalate; MEP, monoethyl phthalate; MBzP, monobenzyl phthalate; BPA, bisphenol A; MP, methylparaben; PP, propylparaben; BP, butylparaben.

**Table S2.** Mean differences for the relation between 16 EDCs and AFC estimated using ordinary least squares regression with models stratified by age in 775 Women in the EARTH Study, 2004-2019. These mean differences can generally be interpreted as the mean difference in AFC for a single unit increase in chemical each chemical for each age group stratum.

| Environmental Chemicals | Conditional<sup>a</sup> Mean Difference (SE<sup>c</sup>) |  | Unconditional<sup>b</sup> Mean Difference (SE<sup>c</sup>) |  |
| --- | --- | --- | --- | --- |
|  | Age < 35 | Age ≥ 35 | Age < 35 | Age ≥ 35 |
| MBP(μg/L) | -1.47 (0.71) | 0.22 (0.50) | -0.20 (0.40) | -0.05 (0.27) |
| MiBP(μg/L) | 1.08 (0.78) | -0.19 (0.42) | 0.47 (0.45) | -0.15 (0.28) |
| MCNP(μg/L) | -0.26 (0.62) | 0.04 (0.38) | 0.08 (0.39) | -0.14 (0.23) |
| MCOP (μg/L) | 0.97 (0.62) | -0.19 (0.37) | 0.49 (0.32) | -0.15 (0.21) |
| MECPP (μg/L) | -1.06 (1.61) | -0.67 (0.72) | -0.38 (0.38) | -0.46 (0.26) |
| MEHHP (μg/L) | 0.83 (2.23) | -0.25 (1.27) | -0.27 (0.35) | -0.37 (0.24) |
| MEHP (μg/L) | 1.08 (0.65) | -0.36 (0.42) | 0.22 (0.36) | -0.45 (0.22) |
| MEOHP (μg/L) | -0.84 (2.73) | 0.64 (1.33) | -0.29 (0.36) | -0.35 (0.24) |
| M CPP (μg/L) | -0.49 (0.81) | 0.17 (0.34) | 0.22 (0.39) | -0.07 (0.21) |
| MEP (μg/L) | 0.08 (0.45) | -0.29 (0.29) | 0.09 (0.38) | -0.22 (0.21) |
| MBzP (μg/L) | 0.59 (0.51) | -0.05 (0.38) | 0.35 (0.39) | -0.03 (0.27) |
| BPA (μg/L) | 0.19 (0.59) | 0.5 (0.34) | -0.01 (0.53) | 0.26 (0.29) |
| BP (μg/L) | -0.31 (0.32) | 0.21 (0.17) | -0.07 (0.25) | 0.08 (0.15) |
| MP (μg/L) | 0.58 (0.75) | 0.29 (0.46) | 0.3 (0.32) | -0.04 (0.19) |
| PP (μg/L) | -0.04 (0.58) | -0.32 (0.35) | 0.21 (0.25) | -0.12 (0.15) |
| Hg (ppm) | -0.72 (0.39) | -0.04 (0.28) | -0.73 (0.37) | 0.02 (0.26) |

a Conditional mean differences obtained from a model where AFC was regressed against all EDC/Hg collectively with an intercept term, stratified by age.
b Unconditional mean differences obtained from a model where AFC was regressed against each EDC/Hg separately with only an intercept and a term for the specific EDC/Hg, stratified by age.
c Standard errors obtained via the HC3 robust variance estimator

